# Electronic Media and its Application in Psychotherapy

**DOI:** 10.64898/2026.02.11.26346102

**Authors:** Ipsit V. Vahia, Julia Kimball, Boyu Ren, Hailey V. Cray, Rebecca Dickinson, Heejung Kim, Dylan Guan, Kerry Ressler

## Abstract

**Background:** As digital communication becomes central to daily life, psychotherapy increasingly has access to patients electronic media data. While digital phenotyping has been widely studied, less is known about whether incorporating personal communication data, such as text messages, improves clinical outcomes in psychotherapy.

**Objective:** To determine whether integrating personalized text message data into psychotherapy improves depression, anxiety, health related quality of life, and therapeutic alliance, and to examine whether increased access to collateral information influences clinical decision making.

**Methods:** The Electronic Media and its Impact on Psychotherapy EMAP study was a randomized controlled trial conducted in adult and geriatric outpatient and partial hospitalization settings at an academic psychiatric hospital. Participants receiving psychotherapy for primary depression or anxiety N = 101 were randomized to Electronic Media Enhanced Therapy EME n = 52 or Treatment as Usual TAU n = 49. In EME, research staff reviewed participants text messages prior to sessions using personalized mood related key terms and presented aggregated findings on a HIPAA compliant clinician dashboard. TAU participants received standard care without message review. Outcomes included change scores for PHQ 9, GAD 7, SF 36, and Working Alliance Inventory Short Revised WAI SR. Linear regression and nonparametric tests compared groups. Moderation analyses assessed whether frequency of information access influenced treatment changes.

**Results:** No significant between group differences were observed in anxiety or working alliance. Among participants with SF 36 follow up data n = 65 EME participants demonstrated greater improvement in the pain subscale. Greater frequency of information access was associated with increased treatment related changes in the EME group. Access to additional collateral information was associated with more clinical actions but not improved distal outcomes.

**Conclusions:** Integrating personalized text message data into short term psychotherapy did not significantly improve depression or anxiety, and neither improved nor worsened therapeutic alliance.

However, access to greater amounts of collateral information including electronic media influenced clinical decision making, suggesting nuanced and context dependent effects.

## Introduction

Over the past decade, the digitization of everyday life has meant that devices such as smartphones serve as conduits for everyday function and communication (1). With interpersonal communication now including digital media such as text messages, blogs, emails, social media, etc., much of this communication is chronicled and preserved in ways that did not exist until the advent of these technologies. Increasingly, psychiatry, and particularly, the realm of psychotherapy, have begun to recognize the ways in which digital data can be incorporated into care. A vast literature exists in the domain of digital biomarkers and digital phenotyping (2-5) where sensors and devices track behavior and, in turn, generate objective measures of activity, sleep, mood and parameters such as medication responses and side effects. However, a simpler and more intuitive use of this digital collateral information that is available through digital media and communication - such as text messages - has been relatively understudied.

Psychotherapy relies on patient recall of key information, events, and interactions. There is evidence to indicate that clinicians sporadically incorporate information from digital communications into therapy sessions, and that including this information may have a positive impact on the psychotherapeutic process (6-8). However, emerging evidence also indicates that the effectiveness of these approaches may be situational and there may be heterogeneity in outcomes, depending on how data are presented and accessed. For instance, one study (9) found that the use of a dashboard that presented clinicians with broad metrics (amount of time spent on their smartphone during the night, miles walked per week, top searches on social media, etc.) led to no statically significant changes in patients’ wellbeing scores. A follow-up study (10) also found that clinician use of such a digital dashboard did improve wellbeing. Notably, both studies found that patients and clinicians alike appreciated how much awareness the dashboard brought to the patients’ digital use. The patients also reported feeling comfortable and positive about the dashboard experience (10). This suggests that the impact of this approach may be subtle and nuanced, a notion that is being increasingly replicated in other studies (11). However, neither study investigated if there was differential impact from evaluating more personal communication such as email or text messages. This is prevalent as Liu et al. (12) found text messages strongly correlate with reported depressive symptoms.

There are important implications to gaining a more sophisticated understanding of how access to broader information from digital communication and media impacts outcomes in psychotherapy. Spending time in a session reviewing digital content can take away from time spent engaged directly with patients (13) and is not clear how this may impact rapport. It is also not clear whether any benefits are empirical or situational. Finally, because there is great inter-individual variance in how individuals across the age span express emotion in electronic communication (14), there is a need for guidelines around how to optimize the use of electronic media. To address these knowledge gaps, we conducted a randomized controlled trial of an intervention where research staff performed a personalized review of patients’ text message communication prior to a psychotherapy session. This information was aggregated into an interactive dashboard for clinicians to access during their session with the patient. We sought to assess whether (a) access to collateral information about text communication impacted depression and anxiety, (b) whether using a dashboard to review this information during therapy sessions impacted the therapeutic alliance, and (c) whether accessing more collateral information (including digital communication) during therapy sessions impacted how therapists’ treatment plan for each session. The Electronic Media and its Impact on Psychotherapy (EMAP)study, was registered on clinicaltrials.gov (NCT03712267) and approved by the Mass General Brigham Institutional Review Board.

We hypothesized that patients receiving care incorporating electronic media would experience greater reductions in depression, anxiety, and improved health-related quality of life scores, as well as a stronger working alliance, compared to those receiving treatment as usual. Additionally, we explored how clinicians used data during sessions and whether increased data access influenced their approach to care

## Methods

### Participants

This study was conducted in the adult and geriatric outpatient and partial hospitalization programs at McLean Hospital, an academic psychiatric hospital affiliated with Mass General Brigham Health and Harvard Medical School. We initially contacted clinicians providing outpatient psychotherapy including psychiatrists, psychologists, and social workers. Clinicians were eligible if they offered psychotherapy, regardless of whether they also prescribed medications concomitantly. Based on referrals from clinicians, we recruited individuals engaged in individual psychotherapy for primary diagnoses of anxiety or depression. Individuals with cognitive impairment, active psychosis, or active substance use disorders were excluded.

Participants were compensated $25 per study visit for their time spent with research assistants reviewing messaging content or completing questionnaires. Study clinicians were compensated $15 per session for the additional time required to complete the study-related questionnaires.

### Intervention

Participants in this study were randomized into two groups: the Electronic Media Enhanced Therapy (EME) group and the Treatment as Usual (TAU) group. The EME group received therapy augmented by the collection and analysis of electronic media data, while the TAU group received standard clinical treatment without the incorporation of electronic media data. Both groups were administered standardized scales. For both the EME and TAU groups, research assistants met with participants prior to each scheduled clinical session. During these meetings, which lasted approximately 30 minutes, study staff administered questionnaires assessing participants’ usage of messaging applications (e.g., text messages, Facebook Messenger, WhatsApp). The questionnaires included both objective measures (e.g., frequency and number of active conversations) and subjective items assessing participants’ comfort with the research process.

In the EME group, research assistants also reviewed participants’ messaging content using Zoom’s screen-sharing feature. Participants were instructed on how to share their screens and were asked to search their messages for 12 pre-selected key terms which reflected emotional valiance for mood states such as depression and anxiety. These terms were chosen based on a consensus of the research team, informed by both clinical experience and prior research. In consultation with the participant’s clinician, each participant was provided with a personalized list of 12 key terms, which were updated as necessary. For participants in partial hospitalization programs, key terms were identified within the previous day’s messaging to account for the shorter duration between clinical visits. The standardized scales administered to both groups were as follows: (a) The Patient Health Questionnaire (9-item) (PHQ-9) (15), (b) the Generalized Anxiety Disorder (7-item) (GAD7) (16), (c). The Working Alliance Inventory – Sort Revised (WAI-SR) (17), (d) The MOS=RAND SF-36 item version (SF-36) (18), (e) The McLean Collateral Information and Clinical Actionability Scale (M-CICAS) (7). Collected data were entered into a custom-designed HIPAA-compliant platform developed by Mirah, a Boston-based company. The Mirah platform included a data collection portal, data storage, and a clinician-facing dashboard that presented the aggregated data from messages. Clinicians reviewed the data before or during psychotherapy sessions. Any content that raised clinical concerns during pre-session data collection was forwarded to the PI and the participant’s therapist for further evaluation and action.

In the TAU group, no review of messaging content occurred. Instead, research assistants focused on collecting self-reported data on mood and health functioning, which were also tracked using the same Mirah platform.

### Research Assistant Training

Before engaging in data collection, all research assistants underwent training to ensure inter-rater reliability. The training, conducted by the principal investigator (PI) or a clinical co-investigator, involved a certification process requiring research assistants to identify 10 emotionally charged sentences out of 20 conversational samples with at least 90% accuracy. Research assistants were also trained to detect content that might raise safety concerns and to follow appropriate procedures for managing these concerns. Once certified, research assistants were authorized to record relevant data, including the frequency of key terms used in participants’ messaging, the emotional tone of these messages, and the identities of the individuals involved in the conversations.

### Assessments

During therapy sessions, clinicians in both groups reviewed the mood scales, health data, and collected collateral information, including messaging behavior for participants in the EME group. Clinicians were encouraged to incorporate this information into their clinical decision-making but retained discretion on how to utilize the data. In the EME group, clinicians could choose to view specific messaging content, especially if there were concerns about suicidality, mood changes, or disorganized thoughts. Any emergent concerns related to safety or mental health were immediately triaged by the therapist or a licensed clinician on the study team, with the PI available for further consultation, if necessary.

After each clinical session, both patients and clinicians completed assessments. The Working Alliance Inventory (WAI) was administered to clinicians and patients at baseline and final visits to assess the therapeutic alliance.

For participants in partial hospitalization programs, the assessment schedule was adjusted to account for their shorter treatment duration. In these cases, mood and health questionnaires were administered between daily clinical sessions.

### Data Collection and Safety Procedures

All collected data, including messaging content and mood assessments, were securely stored on the Mirah platform. Participants retained full autonomy l over access to their devices. Research staff did not have access to participants’ passwords or other security credentials, and participants determined which messages were shared. Any decision to withhold specific messages was respected and formally documented.

When study staff or clinicians identified content suggesting potential safety concerns (e.g., indications of suicidality, psychosis, or marked mood changes), these concerns were promptly escalated to the participant’s therapist and the study principal investigator for immediate evaluation and intervention. The study complied with all applicable mandated reporting requirements and any adverse events were reported to the Institutional Review Board (IRB).

### Statistical Analysis

Demographic characteristics were summarized using means, standard deviations (SD), and ranges for continuous variables and frequency/proportions for categorical variables. Treatment effects were evaluated using change scores for each outcome (SF-36, WAI-SR, PHQ-9, and GAD-7), calculated as the difference between scores at the last session and the first session. SF-36 subscale change scores were modelled as a function of treatment group (EME intervention vs TAU control) using linear regression, controlling for the total number of sessions. This approach was repeated for the PHQ-9 and GAD-7 outcome measurements. Because WAI-SR change scores violated normality assumptions, Wilcoxon rank-sum tests were used for this outcome.

Secondary analyses were conducted within the subsample of participants enrolled in the partial hospitalization program. Finally, moderation analyses were performed to determine whether the treatment group (EME vs. TAU) modified the association between frequency of information access and treatment changes using the M-CICAS measures. Given the small sample size, no adjustments for multiple comparisons were applied. Statistical significance was defined as p < 0.05 for all tests. All analyses were conducted using SPSS version 26.0.

## Results

The study included 101 participants, with 52 assigned to the EME intervention group and 49 to the TAU control group. Most participants (86.1%) were enrolled in a partial hospitalization program. Across the cohort, the mean age was 39.4 years (SD = 6.2), 63.4% identified as female, and 80.2% self-identified as White [Table 1]. Participants reported an average of 16.3 ± 2.3 years of education. The most common primary psychiatric diagnoses were depression (74.3%) and anxiety (21.8%). No significant differences were observed between the EME and TAU groups across demographic variables.

**Table 1.** Sample characteristics at baseline.

| Variable | Total | EME | TAU |
| --- | --- | --- | --- |
| Age (years) | 39.4 (16.2), 19-80 | 40.6 (16.3), 19-70 | 38.2 (16.2), 19-80 |
| Sex |  |  |  |
| Female | 64 (63.4) | 30 (57.7) | 34 (69.4) |
| Male | 25 (24.8) | 17 (32.7) | 8 (16.3) |
| Other | 4 (4.0) | 2 (3.8) | 2 (4.1) |
| Missing | 8 (7.9) | 3 (5.8) | 5 (10.2) |
| Education (years) | 16.3 (2.3), 12-22 | 16.7 (2.39), 12-22 | 15.9 (2.0), 12-20 |
| Race |  |  |  |
| White | 81 (80.2) | 43 (82.7) | 38 (77.6) |
| Black or African American | 3 (3.0) | 0 (0) | 3 (6.1) |
| Asian | 5 (5.0) | 3 (5.8) | 2 (4.1) |
| Other | 8 (7.9) | 6 (11.5) | 2 (4.1) |
| Missing | 4 (4.0) | 0 (0) | 4 (8.2) |
| Diagnosis |  |  |  |
| Depression | 75 (74.3) | 38 (73.1) | 37 (75.5) |
| Anxiety | 22 (21.8) | 13 (25.0) | 9 (18.4) |
| Missing | 4 (4.0) | 1 (1.9) | 3 (6.1) |

*Note.* Numeric variables are presented in mean (standard deviation), range. Categorical variables are presented in n (%). All table values were rounded to one decimal place.

Participants in the EME group exhibited greater reductions in PHQ-9 and GAD-7 scores from their first and last session compared to those in the TAU group; however, these differences were not statistically significant (PHQ-9 *b* = -0.62, 95% CI: [-2.87, 1.62], *p* = .58; GAD-7 *b* = -0.26, 95% CI: [-1.90, 1.39], *p* =.75). Among the subset of 65 participants with follow-up SF-36 data, no significant differences were observed between groups across SF-36 domains, except for pain [Table 2]: EME participants demonstrated a 9.54-point greater increase in SF-36 pain subscale scores compared to TAU participants (95% CI: [1.94, 17.14], *p* = .02). This effect persisted within the subsample enrolled in the partial hospitalization program (*b* = 8.55, 95%CI: [0.55, 16.55], *p* =.04).

**Table 2.** Change in SF-36 subscales between first and last session across EME and TAU groups.

| SF-36 subscale | b | 95% CI | p |
| --- | --- | --- | --- |
| Full sample |  |  |  |
| Emotional Wellbeing | -2.09 | -8.60, 4.42 | .53 |
| Energy/Fatigue | 3.77 | -3.19, 10.73 | .29 |
| General Health | 0.27 | -5.45, 5.99 | .93 |
| Pain | 9.54 | 1.94, 17.14 | .02 |
| Physical Functioning | 0.70 | -4.65, 6.05 | .80 |
| Role Limitations |  |  |  |
| Emotional | 0.71 | -9.52, 10.94 | .89 |
| Physical | 1.50 | -12.49, 15.49 | .83 |
| Social Functioning | 2.42 | -7.01, 11.85 | .62 |
| Partial program |  |  |  |
| Emotional Wellbeing | -3.27 | -10.13, 3.59 | .35 |
| Energy/Fatigue | 5.18 | -2.09, 12.45 | .17 |
| General Health | 0.10 | -5.56, 5.76 | .97 |
| Pain | 8.55 | 0.55, 16.55 | .04 |
| Physical Functioning | 2.11 | -3.40, 7.62 | .46 |
| Role Limitations |  |  |  |
| Emotional | 1.76 | -9.29, 12.81 | .76 |
| Physical | 3.98 | -10.66, 18.62 | .60 |
| Social Functioning | 1.40 | -8.60, 11.40 | .79 |

*Note.* b coefficients were estimated from linear regression in the total sample with follow-up SF-36 data (n=65), as well as the subgroup of participants in the partial program (n=60). Models were adjusted for the number of total sessions received. All table values were rounded to two decimal places.

Wilcoxon rank-sum tests further revealed that there were no group differences in change in WAI-SR item scores [Table 3] among participants with available follow-up data (n=41).

**Table 3.** Change in WAI-SR item scores between first and last session across EME and TAU groups.

| WAI-SR item | Mean Diff. | p |
| --- | --- | --- |
| Full sample |  |  |
| 1. As a result of these sessions I am clearer as to how I might be able to change. | -0.39 | .26 |
| 2. What I am doing in therapy gives me new ways of looking at my problem | -0.13 | .61 |
| 3. I believe___likes me. | 0.24 | .10 |
| 4. ___and I collaborate on setting goals for my therapy. | 0.03 | .47 |
| 5. ___and I respect each other. | 0.04 | .45 |
| 6. ___and I are working towards mutually agreed upon goals. | 0.10 | .74 |
| 7. I feel that___appreciates me. | -0.26 | .38 |
| 8. _____ and I agree on what is important for me to work on. | 0.09 | .81 |
| 9. I feel _____ cares about me even when I do things that he/she does not approve of. | -0.04 | .91 |
| 10. I feel that the things I do in therapy will help me to accomplish the changes that I want. | 0.32 | .26 |
| 11. _____ and I have established a good understanding of the kind of changes that would be good for me. | -0.29 | .17 |
| 12. I believe the way we are working with my problem is correct. | 0.10 | .99 |
| Partial program |  |  |
| 1. As a result of these sessions I am clearer as to how I might be able to change. | -0.43 | .21 |
| 2. What I am doing in therapy gives me new ways of looking at my problem | -0.09 | .75 |
| 3. I believe___likes me. | 0.25 | .10 |
| 4. ___and I collaborate on setting goals for my therapy. | 0.10 | .30 |
| 5. ___and I respect each other. | 0.04 | .45 |
| 6. ___and I are working towards mutually agreed upon goals. | 0.26 | .40 |
| 7. I feel that___appreciates me. | -0.23 | .50 |
| 8. _____ and I agree on what is important for me to work on. | 0.14 | .63 |
| 9. I feel _____ cares about me even when I do things that he/she does not approve of. | -0.09 | .77 |
| 10. I feel that the things I do in therapy will help me to accomplish the changes that I want. | 0.43 | .15 |
| 11. _____ and I have established a good understanding of the kind of changes that would be good for me. | -0.22 | .25 |
| 12. I believe the way we are working with my problem is correct. | 0.16 | .82 |

*Note.* The number of participants with follow-up WAI-SR data is n=41. The subgroup of participants in the partial program is n = 39. All table values were rounded to two decimal places.

The association between treatment changes and the frequency of information access differed by assignment to EME or TAU arms (*p* = .02). Specifically, treatment changes were positively associated with the number of times information was accessed in the EME group (*b* = 0.72) but not in the TAU group (*b* = -0.01, 95% CI: [0.12, 1.34], *p* = .02) (Figure 1).

**Figure 1.**
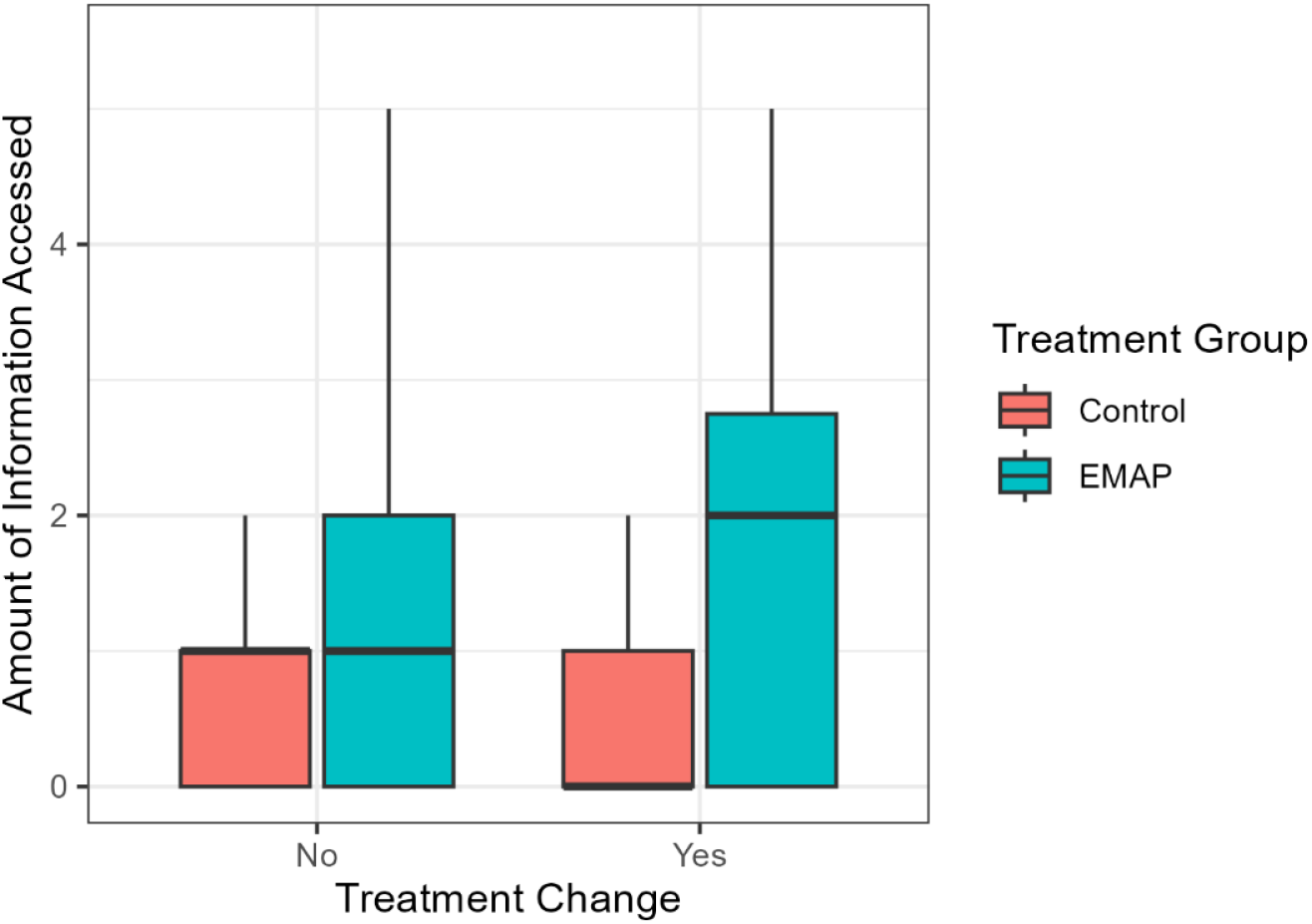
Distribution of the number of sources of information accessed by treatment group and treatment change status.

## Discussion

This study examined whether incorporating data from electronic media into psychotherapy influences clinical outcomes and the therapeutic alliance between clinicians and patients. It also explored how integrating data from multiple sources informs clinical decision-making. At a time when there are ever-growing streams of data becoming accessible to clinicians, this study addresses the fundamental question of whether and how these data can add value. In this study of 101 clinician-patient dyads, no statistically significant differences were observed in PHQ-9 or GAD-7 scores between the EME and TAU groups, suggesting that incorporating electronic media did not improve distal clinical outcomes related to anxiety and depression. Similarly, Working Alliance Inventory (WAI) scores not differ between groups, indicating that the therapeutic alliance was unaffected by the inclusion of electronic media. Consequently, the study’s primary hypotheses were not supported.

, Exploratory analyses indicated that accessing a greater number of information sources was associated with an increased number of clinical actions taken during visits. This finding encompasses all collateral information sources, not solely digital data. . However, taking more clinical actions did not translate into improved outcomes when comparing groups. Nonetheless, this exploratory finding suggests that clinicians tend to make more changes during psychotherapy when provided with additional collateral information. One possible reason for the lack of support for the primary hypothesis may relate to characteristics of the study sample, as most participants were enrolled in a partial hospitalization program and received care only for two weeks, during which we gathered data for an average of fewer than four sessions per participant. This may simply not have been sufficient contact to capture change in the downstream variables. However, one may anticipate that on the individual level, there may be benefits to more clinical actions from accessing more data that are not captured by our selected outcome variables. These findings are consistent with existing studies (9, 10) which provided clinicians with access to a dashboard that displayed data from their social media and Internet use. These studies found that while providing clinicians with this dashboard was feasible and that responses to access to this dashboard were generally positive, distal clinical outcomes did not improve. Our finding also provides a counter point to an extant literature that has suggested that access to informatics may universally improve outcomes (19).

Several limitations of this study should be noted. The relatively small sample size, which primarily included participants in a short-term outpatient service is a key factor, since it meant that we were only able to track participants for a maximum of two weeks. While our study included participants who were engaged in ongoing outpatient care, their number may have been too small to detect clinically significant effects. Additionally, the study was impacted by the COVID-19 pandemic. Originally designed and funded for in-person therapy, the protocol was adapted to include virtual sessions. The extent to which this transition influenced study outcomes remains unclear, and despite protocol adjustments, unmeasured variables may have affected results.

This project highlights several meaningful directions for future research. Chief among these is that there may be subtleties around the ways in which accessing a person’s electronic media can impact their psychotherapy and these subtleties are likely specific to each individual. Although the primary hypotheses were not supported, the exploratory findings suggest that providing clinicians with additional collateral information, including digital data, can influence clinical decision-making (20). However, broad outcomes such as improvement in depression or anxiety, particularly in a study where participants were tracked for as few as two weeks, may not capture this impact. We recommend that future work incorporate novel designs that can capture intra-individual changes in response to access to electronic media for psychotherapy.

## Data Availability

All data produced in the present study are available upon reasonable request to the authors

## Acknowledgements

Patrick Monette, Katherine Hobbs, Rose May, Aniqa Rahman, Miranda Skurla, Praise Owoyemi, Benjamin Silverman, Courtney Beard, and Laura Germine, The clinicaltrials.gov identifying number for this study is NCT03712267

